# Breadth of concomitant immune responses underpinning viral clearance and patient recovery in a non-severe case of COVID-19

**DOI:** 10.1101/2020.02.20.20025841

**Authors:** Irani Thevarajan, Thi HO Nguyen, Marios Koutsakos, Julian Druce, Leon Caly, Carolien E van de Sandt, Xiaoxiao Jia, Suellen Nicholson, Mike Catton, Benjamin Cowie, Steven YC Tong, Sharon R Lewin, Katherine Kedzierska

## Abstract

We report the kinetics of the immune response in relation to clinical and virological features of a patient with mild-to-moderate coronavirus disease-19 (COVID-19) requiring hospitalisation. Increased antibody-secreting cells, follicular T-helper cells, activated CD4^+^ and CD8^+^ T-cells and IgM/IgG SARS-CoV-2-binding antibodies were detected in blood, prior to symptomatic recovery. These immunological changes persisted for at least 7 days following full resolution of symptoms, indicating substantial anti-viral immunity in this non-severe COVID-19.

---

On 30/01/2020, a 47-year old woman from Wuhan, Hubei province, China presented to an emergency department in Melbourne, Australia. Her symptoms commenced 4 days earlier with lethargy, sore throat, dry cough, pleuritic chest pain, mild dyspnoea and subjective fevers (**Fig.1a)**. She had travelled 11 days prior to presentation from Wuhan via Guangzhou to Australia. She had no contact with the Huanan seafood market or known COVID-19 cases. She was otherwise healthy, non-smoker, taking no medications. Clinical examination revealed a temperature of 38.5°C, pulse rate 120 beats/minute, blood pressure 140/80 mmHg, respiratory rate 22 breaths/minute, and oxygen saturation 98%, while breathing ambient air. Lung auscultation revealed bibasal rhonchi. At presentation day (d) 4, SARS-CoV-2 was detected by real-time reverse transcriptase polymerase-chain-reaction (rRT-PCR) from a nasopharyngeal swab specimen. SARS-CoV-2 was again detected at d5-6 from nasopharyneal, sputum and faecal samples, but was undetectable from d7 (**Fg**.**1a**). Blood C-reactive protein was elevated at 83.2, with normal lymphocyte counts (4.3×10^9^/L [range 4.0-12.0×10^9^/L]) and normal neutrophil counts (6.3×10^9^/L [range 2.0-8.0×10^9^/L]). No other respiratory pathogens were detected. Her management was intravenous fluid rehydration without supplemental oxygenation. No antibiotics, steroids or antiviral agents were administered. Chest radiography demonstrated bibasal infiltrates at d5, which cleared on d10 (**Fig.1b)**, and she was discharged to home isolation on d11. Symptoms resolved completely by d13 and she remained well at d20 post-onset of symptoms. Progressive increase in plasma COVID-19-binding IgM/IgG antibodies, from d7 until d20 was observed **(Fig.1c)**.

**Fig.1.**
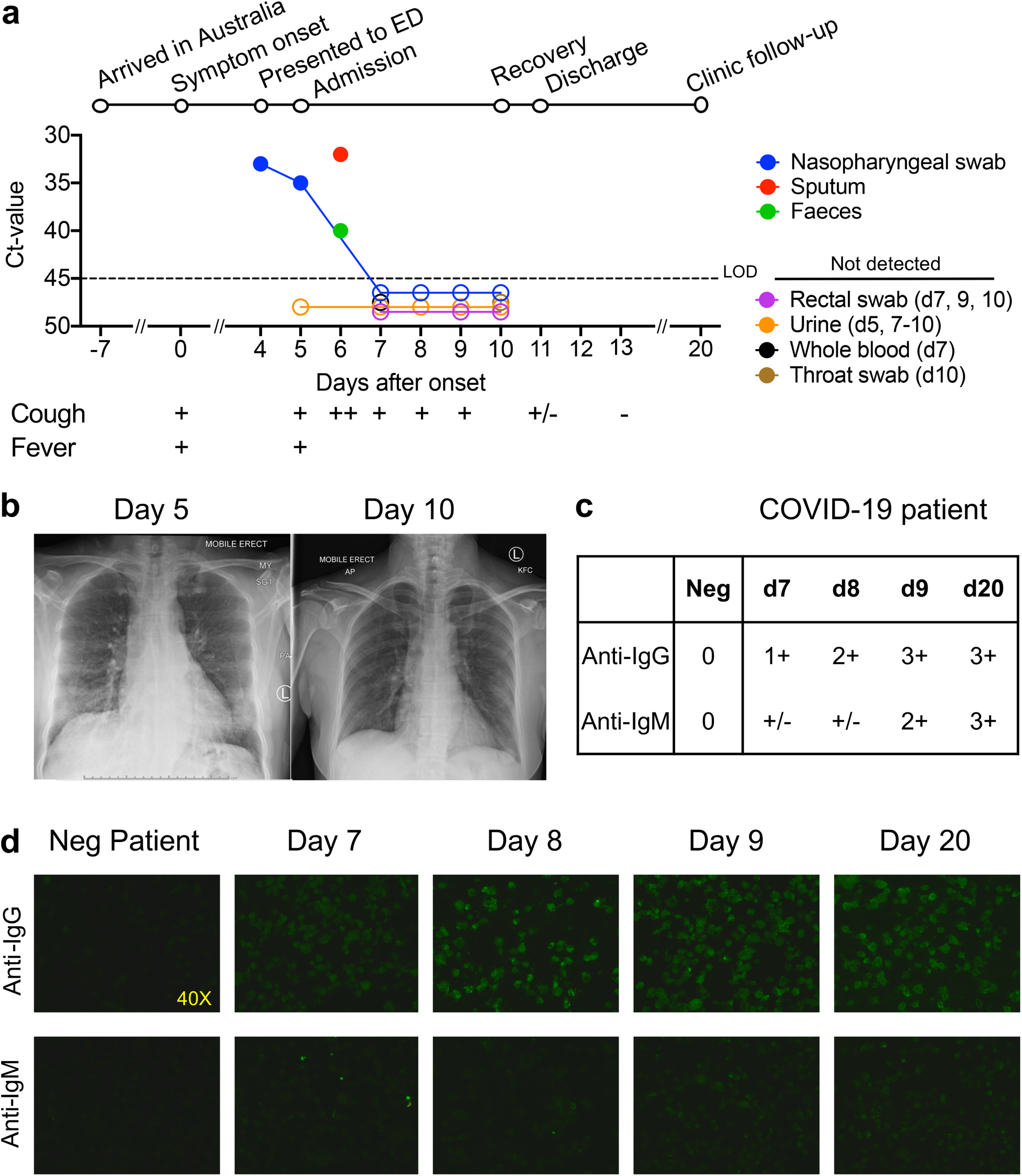
Time course of clinical presentation and detection of SARS-CoV-2 in a range of clinical specimens and antibodies to SARS-CoV2 in blood. **(a)** Timeline of COVID-19; detection of SARS-CoV-2 virus in sputum, nasopharyngeal aspirates, faeces but not urine, rectal swab and whole blood. SARS-CoV-2 was quantified by real-time RT-PCR and the cycle threshold (Ct) is shown for each of the specimen. An increase in Ct value is consistent with a decrease in viral load. The assay limit of detection (LOD) threshold is Ct=45. Open circles: undetectable SARS-CoV-2; **(b)** Radiological improvement from admission to discharge from hospital. Anteroposterior chest radiographs on d5 (day of admission) and d10 following onset of symptoms; **(c, d)** Immunofluorescence antibody staining for the detection of IgG and IgM bound to SARS-CoV-2-infected vero cells using plasma (diluted 1:20) collected at d7-9 and d20 following onset of symptoms.

There are currently no data defining immune responses leading to viral clearance and clinical resolution of COVID-19. We addressed this knowledge gap by analysing the breadth of immune responses in blood prior to patient recovery. As antibody-secreting cells (ASCs) are key for the rapid production of antibodies following viral Ebola infection^1,2^, influenza virus infection and vaccination^2,3^; and activated circulating follicular T helper (cTfhs) are concomitantly induced following influenza vaccination^3^, we first determined the frequency of CD3^-^CD19^+^CD27^hi^CD38^hi^ ASCs and CD4^+^CXCR5^+^ICOS^+^PD1^+^ cTfh responses at 3 days prior to symptomatic recovery. ASCs appeared in blood at the time of viral clearance at d7 (1.48%) and peaked on d8 (6.91%). Emergence of cTfhs occurred concurrently in blood at d7 (1.98%), with the frequency increasing on d8 (3.25%) and d9 (4.46%) (**Fig.2a**). The peak of both ASCs and cTfhs was markedly higher in the COVID-19 patient than the baseline levels in healthy controls (average±SD: 0.61±0.40% and 1.83±0.77%, respectively, n=5). Both ASCs and cTfhs were still prominently present at convalescence (d20) (4.54% and 7.14%, respectively; **Fig.2a**). Our study provides evidence on the recruitment of both ASCs and cTfhs in patient’s blood whilst still unwell and 3 days prior to resolution of symptoms, indicating their importance in anti-viral immunity towards SARS-CoV-2.

**Fig.2.**
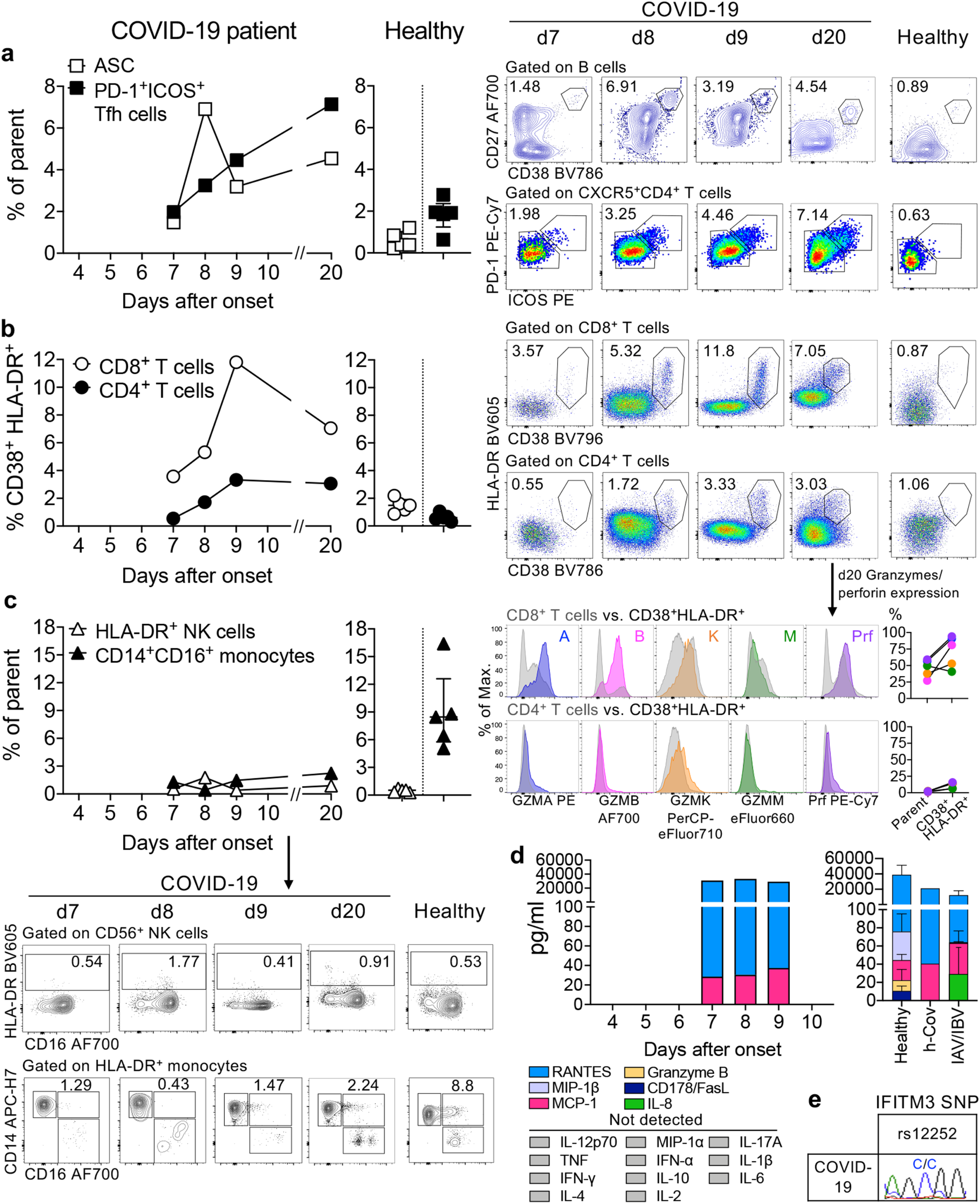
Emergence of immune responses during non-severe symptomatic COVID-19. Frequencies of **(a)** CD27^hi^CD38^hi^ antibody-secreting cells (ASC; plasmablasts, gated on CD3^-^CD19^+^ lymphocytes) and activated ICOS^+^PD1^+^ follicular T helper (Tfh) cells (gated on CD4^+^CXCR5^+^ lymphocytes); **(b)** activated CD38^+^HLA-DR^+^ CD8^+^ and CD4^+^ T-cells; **(c)** lineage^-^CD14^+^CD16^+^ monocytes and activated HLA-DR^+^ NK cells (gated on CD3^-^CD14^-^CD56^+^ cells) detected by flow cytometry for blood collected at d7-d9 and d20 following onset of symptoms and in healthy donors (median with IQ range); **(b)** Histograms and line graphs of granzymes A/B/K/M and perforin (Prf) staining of parent CD8^+^ or CD4^+^ T cells and activated CD38^+^HLA-DR^+^CD8^+^/CD4^+^ T-cells are shown (bottom panels). **(d)** Plasma levels of pro-inflammatory cytokines/chemokines in COVID-19 patient at d7-9, healthy individuals (n=5, mean±SEM), patient with HCoV-229e and influenza-infected patients (n=5). **(e)** ‘risk’ IFITM3-rs12252 genotyping for the COVID-19 patient.

Since co-expression of CD38 and HLA-DR is well defined as the key phenotype of CD8^+^ T-cell activation towards viral infections, we analyzed activation of CD8^+^ T-cells by CD38/HLA-DR co-expression. In accordance with previous reports on Ebola and influenza^1,4^, CD38^+^HLA-DR^+^ co-expression on CD8^+^ T-cells rapidly increased from d7 (3.57%) to d8 (5.32%) and d9 (11.8%), with a decrease at d20 (7.05%) (**Fig.2b**). Furthermore, the frequency of CD38^+^HLA-DR^+^ co-expression on CD8^+^ T-cells in this patient was markedly higher than on CD8^+^ T-cells in healthy individuals (1.47±0.50%, n=5). Similarly, CD38^+^HLA-DR^+^ co-expression increased on CD4^+^ T-cells between d7 (0.55%) and d9 (3.33%) in the patient, compared to healthy donors (0.63±0.28%, n=5), although at lower levels than CD8^+^ T-cells. CD38^+^HLA-DR^+^ T cells, especially within CD8^+^ T-cells, produced higher amounts of granzymes A/B and perforin (∼34-54% higher) than their parent (CD8^+^ or CD4^+^ populations, **Fig.2b**). Thus, the emergence and rapid increase in activated CD38^+^HLA-DR^+^ T-cells, especially CD8^+^ T-cells, at d7-9 preceded resolution of symptoms.

We also analysed CD16^+^CD14^+^ monocytes, related to immunopathology, and activated HLA-DR^+^CD3^-^CD56^+^ NK cells (**Fig.2c**). We detected reduced frequencies of CD16^+^CD14^+^ monocytes in peripheral blood at d7-9 (1.29%, 0.43%, 1.47%, respectively), compared to healthy controls (9.03±4.39%, n=5). This might indicate efflux of CD16^+^CD14^+^ monocytes from blood to the site of infection, which remained low at d20 (2.24%). Low levels of activated HLA-DR^+^CD3^-^CD56^+^ NK cells were found in both the COVID-19 patient and healthy controls.

As high levels of pro-inflammatory cytokines/chemokines are predictive of severe clinical outcomes for influenza^5^, seventeen pro-inflammatory cytokines/chemokines were quantified in patient’s plasma. We found low levels of monocyte chemoattractant protein-1 (MCP-1; CCL2), important for the recruitment of monocytes, T-cells and dendritic cells to the site of infection (**Fig.2d**). However, these MCP-1 levels were similar to healthy donors (22.15±13.81, n=5), patients infected with influenza A (IAV) and influenza B viruses (IBV) at d7-9 (33.85±30.12, n=5) and a patient with a known human coronavirus infection HCoV-229e (hCoV, 40.56). Substantial levels of RANTES (CCL5), involved in homing and migration of activated T-cells that express CCR5, were also detected in COVID-19 plasma but these were comparable to healthy donors (p=0.412), IAV/IBV-infected patients (p=0.310) and a hCoV-patient. Thus, in contrast to severe avian H7N9 disease with highly elevated IL-6 and IL-8, and intermediate IL-10, MIP-1β, IFN-γ^5^, minimal pro-inflammatory cytokines/chemokines were found in this patient with CoVID-19, even while symptomatic at d7-9.

Given that interferon-induced transmembrane protein-3 (IFITM3) single nucleotide polymorphism (SNP)-rs12252-C/C was linked to severe influenza^5,6^, we analysed the IFITM3-rs12252 SNP in this patient with COVID-19. Interestingly, the patient had the ‘risk’ IFITM3-rs12252-C/C variant (**Fig.2e**), associated with clinical compromise for 2009-pH1N1^6^ and severe/fatal avian H7N9 disease^5^. As the relative prevalence of IFITM3-rs12252-C/C risk variant in a healthy Chinese population is 26.5% (data from 1,000 genome project)^5^, further investigations of the IFITM3-rs12252-C/C allele in larger cohorts of patients with COVID-19 and its correlation with disease severity is worth pursuing.

Collectively, our study provides novel contributions to the understanding of the breadth of the immune response during a non-severe case of COVID-19. This patient did not experience complications of respiratory failure, acute respiratory distress syndrome, did not require supplemental oxygenation and was discharged within a week of hospitalization, consistent with non-severe but clearly symptomatic disease. We provide evidence on the recruitment of immune populations (antibody-secreting B cells, follicular T-cells, activated CD4^+^ and CD8^+^ T-cells), together with IgM-IgG SARS-CoV-2-binding antibodies, in patient’s blood prior to resolution of clinical symptoms. We propose that these immune parameters should be characterised in larger cohorts of patients with COVID-19 with different disease severity to understand whether they could be used to predict disease outcome and to evaluate new interventions to minimise severity and/or to inform protective vaccine candidates. Furthermore, our study indicates that robust multi-factorial immune responses can be elicited towards the newly-emerged SARS-CoV-2, and similar to the avian H7N9 disease^7^, early adaptive immune responses might correlate with better clinical outcomes.

## Data Availability

Data availability
The data that support the findings of this study are available from the corresponding author upon request. Raw FACS data are shown in the manuscript.

## Acknowledgments

The authors thank Prof Cameron Simmons for supporting the development of SETREP-ID, all the SETREP-ID investigators for their support and Australian Partnership for Preparedness Research for Infectious Disease Emergencies (APPRISE) for ongoing funding of SETREP-ID. We thank Dr Louise Rowntree for technical assistance. This work was funded by the Australian National Health and Medical Research Council (NHMRC) Investigator Grant to KK (#1173871). CES has received funding from the European Union’s Horizon 2020 research and innovation programme under the Marie Skłodowska-Curie grant agreement No 792532 and University of Melbourne McKenzie Fellowship laboratory support. KK is supported by a NHMRC Senior Research Fellowship Level B (#1102792) and SRL is supported by an NHMRC Practitioner Fellowship and an NHMRC program grant. SYCT is supported by a NHMRC Career Development Fellowship (#1145033). XJ is supported by China Scholarship Council-University of Melbourne joint Scholarship. The authors wish to acknowledge our public health partners, and VIDRL’s major funder, the Victorian Department of Health and Human Services without whom this work would not have been possible, and the clinical and laboratory staff involved in the care of this patient.

## Online content

### Author contribution

IT, THON, MK, CES, LC, SN, XJ, JD, MK, BC, SYT, SRL, KK formulated ideas, designed the study and experiments; THON, MK, CES, LC, SN, XJ, JD performed experiments; THON, MK, LC, SN, JD analysed the experimental data, KK, IT, THON, SYT, BC, SRL wrote the manuscript. All authors reviewed the manuscript.

## Methods

### Study design

The patient was enrolled through the Sentinel Travelers Research Preparedness Platform for Emerging Infectious Diseases novel coronavirus substudy (SETREP-ID coV). Sputum, nasopharyngeal aspirates, urine and faecal specimens as well as whole blood in sodium heparin tubes were collected over the duration of illness and 9 days post discharge for quantitative virology, immunology and assessment of host gene factors. Human experimental work was conducted according to the Declaration of Helsinki Principles and according to the Australian National Health and Medical Research Council Code of Practice. Participants provided written informed consent prior to the study. The study was approved by the Royal Melbourne Hospital (HREC Reference number: HREC/17/MH/53 and HREC/15/MonH/64/2016.196) and University of Melbourne (ID #1442952.1 and #1443389.4) Human Research Ethics Committees.

### Generation of SARS-CoV-2 cDNA

RNA was extracted from 200μL from patient’s swabs (nasopharyngeal, rectal, throat in VTM), sputum, urine, faeces and whole-blood samples using the QIAamp 96 Virus QIAcube HT Kit (Qiagen, Hilden, Germany). Reverse transcription was performed using the BioLine SensiFAST cDNA kit (Bioline, London, United Kingdom). Total reaction mixture of 20μl contained 10μL of the RNA extract, 4μl of 5x TransAmp buffer, 1μl of Reverse Transcriptase and 5μl of Nuclease free water. Reactions were incubated at 25°C for 10 min, 42°C for 15 min, 85°C for 5 min.

### Nested SARS-CoV-2 RT-PCR and Sanger sequencing

A PCR mixture containing 2μl cDNA, 1.6μl of 25 mM MgCl_2_, 4μl of 10x Qiagen Taq Buffer, 0.4μl of 20mM dNTPs, 0.3μl of Taq polymerase (Qiagen, Hilden, Germany) and 2μl of 10μM primer pools as described^11^. The first round included the forward (5’-GGKTGGGAYTAYCCKAARTG-3’) and reverse (5’-GGKTGGGAYTAYCCKAARTG-3’) primers. Cycling conditions were 94°C for 10 min, followed by 30 cycles of 94°C for 30s, 48°C for 30s and 72°C for 40s, with a final extension of 72°C for 10 min. PCR product (2μl) was used in the second round PCR reaction which included the forward (5’-GGTTGGGACTATCCTAAGTGTGA-3’) and reverse (5’-CCATCATCAGATAGAATCATCAT-3’) primers. Cycling conditions were 94°C for 10min, followed by 40 cycles of 94°C for 30s, 50°C for 30s and 72°C for 40s, with a final extension of 72°C for 10 min. PCR products had an expected size of approximately 440bp on a 2% agarose gel. The PCR products were purified using ExoSAP-IT (Affymetrix, Santa Clara, CA, USA) and sequenced using an Applied Biosystems SeqStudio Genetic Analyzer (Life Technologies, Carlsbad, CA, USA) using Big Dye Terminator 3.1 (Life Technologies, Carlsbad, CA, USA) and Round 2 PCR primers above. SARS-CoV and 229e-CoV cDNA were used as positive controls.

### Detection of SARS-CoV-2 using TaqMan Real-time RT-PCR E-gene assay

TaqMan RT-PCR assay comprised of 2.5μl cDNA, 10μl Primer Design PrecisonPLUS qPCR Master Mix 1μM Forward (5’-ACA GGT ACG TTA ATA GTT AAT AGC GT −3’), 1μM Reverse (5’-ATA TTG CAG CAG TAC GCA CAC A-3’) primers and 0.2μM Probe (5’-FAM-ACA CTA GCC ATC CTT ACT GCG CTT CG-NFQ-3’) targeting the Betacoronavirus E-gene^1^. The real-time RT-PCR assay was performed on an Applied Biosystems ABI 7500 Fast Real-time PCR machine (Applied Biosystems, Foster City, CA, USA) with cycling conditions 95°C for 2min, 95°C for 5s, 60°C for 24s. SARS-CoV cDNA (Ct∼30) was used as a positive control.

### IFITM3 SNP analysis

PCR was performed on genomic DNA extracted from patient’s granulocytes (using QIAamp DNA Mini Kit, QIAGEN) to amplify the *exon 1* rs12252 region using forward (5′-GGAAACTGTTGAGAAACCGAA-3′) and reverse (5′-CATACGCACCTTCACGGAGT-3′) primers^2^.

### Cytokine analysis

Patient’s plasma was diluted 1:4 before measuring cytokine levels (IL-2, IL-4, IL-6, IL-8, IL-10, IL-12p70, IL-17A, IL-1β, IFN-α, MIP-1α, MIP-1β, MCP-1, CD178/FasL, granzyme B, RANTES, TNF, IFN-γ) using the Human CBA Kit (BD Biosciences, San Jose, California, USA). For RANTES, sera/plasma was also diluted to 1:50. Healthy donors D6-D10 were of a mean age of 32 (range 22-55 years; 40% females).

### Whole blood staining and flow cytometry

Fresh whole blood (200μl per stain) was used to measure CD4^+^CXCR5^+^ICOS^+^PD1^+^ follicular T cells (Tfh) and CD3^-^CD19^+^CD27^hi^CD38^hi^ antibody-secreting B cell (ASC; plasmablast) populations as described^3^ as well as activated HLA-DR^+^CD38^+^CD8^+^ and HLA-DR^+^CD38^+^CD4^+^ T cells, inflammatory CD14^+^CD16^+^ and conventional CD14+ monocytes, activated HLA-DR^+^CD3^-^CD56^+^ NK cells, as per the specific antibody panels (Supplementary Table 1; gating strategy is presented in Supplementary Fig.1). After the whole blood was stained for 20 mins at room temperature (RT) in the dark, samples were lysed with BD FACS Lysing solution, washed and fixed with 1% PFA. Granzymes/perforin staining (patient d20) was performed using the eBioscience Foxp3/Transcription Factor Staining Buffer Set after the lysis step. All the samples were acquired on a LSRII Fortessa (BD). Flow cytometry data were analyzed using FlowJo v10 software. Healthy donors D1-D5 were of a mean age of 35 (range 24-42 years, 40% females).

### Detection of IgG and IgM antibodies in SARS-CoV-2 -infected vero cells

Immunofluorescence antibody tests for the detection of IgG and IgM were performed using SARS-CoV-2-infected vero cells that had been washed with PBS and methanol/acetone fixed onto glass slides. Ten μL of a 1/20 dilution of patient plasma in PBS from days 7, 8, 9 and 20 were incubated on separate wells for 30 mins at 37°C, then washed in PBS and further incubated with 10μL of FITC-conjugated goat anti-human IgG and IgM (Euroimmun, Lűbeck, Germany) before viewing on a EUROStar III Plus fluorescent microscope (Euroimmun). Prior to detection of IgM antibodies, samples were pre-treated with RF-SorboTech (Alere, Rűsselsheim, Germany) to remove IgG antibodies and rheumatoid factors, which may cause false-negative and false-positive IgM results, respectively.

## Data availability

The data that support the findings of this study are available from the corresponding author upon request. Raw FACS data are shown in the manuscript.

---

**Supplementary Table 1. Whole blood immunophenotyping and antibody panels used in our immune assays**.

**Supplementary Figure 1. Flow cytometry gating strategy for immune cell subsets**. Gating panels are shown for **(a)** CD27^hi^CD38^hi^ ASCs and activated ICOS^+^PD1^+^ Tfh cells; **(b)** activated CD38^+^HLA-DR^+^ CD8^+^ and CD4^+^ T-cells, activated HLA-DR^+^ NK cells and lineage^-^CD14^+^CD16^+^ monocytes; and **(c)** granzymes (GZM) A/B/K/M and perforin expression on CD8^+^/CD4^+^ T-cells and activated CD38^+^HLA-DR^+^ CD8^+^/CD4^+^ T-cells.

